# Intergenerational Transmission of Parental Mental Health and Wellbeing Traits on Offspring Emotional and Behavioural Difficulties

**DOI:** 10.1101/2025.06.17.25329751

**Authors:** Robyn E Wootton, Laurie J Hannigan, Leonard Frach, Hannah M Sallis, Elizabeth C Corfield, Andrea G. Allegrini, Amy Shakeshaft, Rebecca M Pearson, Jean-Baptiste Pingault, Ole A. Andreassen, Anita Thapar, Helga Ask, Alexandra Havdahl

## Abstract

**Importance:** Parental mental health and wellbeing are strongly associated with offspring emotional, behavioural and temperament difficulties in the preschool years, which in turn are linked to later mental illness. Understanding how intergenerational transmission of parental mental health and wellbeing traits occurs can inform intervention strategies.

**Objective:** Intergenerational transmission of mental health and wellbeing traits can occur through both genetic and environmental pathways, which can be disentangled using genetic data from mother-father-child trios. In preschool-aged offspring, we estimated the relative importance of “genetic transmission”, where parental genetic factors directly transmitted to the offspring influence offspring traits, *versus* “genetic nurture”, where parental genetic factors shape the child’s environment, which in turn influences the offspring’s traits.

**Design:** We used a trio-polygenic score design. Our primary model used structural equation modelling with full information maximum likelihood estimation to account for missing data.

**Setting:** We used data from the Norwegian Mother Father and Child Cohort Study (MoBa), a pregnancy-based population cohort recruited in Norway from 1999-2008.

**Participants:** The model included up to 43,036 genotyped trios with child outcome measurements.

**Exposures:** Parental genetic predisposition to mental health and wellbeing traits: depression, anxiety, neuroticism, wellbeing, life satisfaction and positive affect.

**Main Outcomes/Measures:** Longitudinal development of mother-rated offspring emotional, behavioural and temperament difficulties from age 1.5- to 5-years.

**Results:** Genetic nurture explained more variance in preschool emotional, behavioural and temperament difficulties than did direct genetic transmission. There was evidence for significantly larger maternal than paternal genetic nurture effects, in particular for positive affect, life satisfaction and anxiety PGS. Genetic nurture from wellbeing traits was associated with reduced preschool emotional, behavioural and temperament difficulties. Genetic nurture effects on behavioural difficulties were significantly larger from wellbeing traits than mental health traits.

**Conclusions:** Our findings suggest a greater relative importance of genetic nurture in the associations between parental mental health and wellbeing traits on preschool emotional, behavioural and temperament difficulties. We found genetic nurture for parental wellbeing traits, consistent with theories that positive family environments could foster resilience in preschool children beyond inherited genetic predispositions. Further investigation to identify specific components of genetic nurture estimates is warranted.

## Introduction

Depression and anxiety are leading causes of global disability^1^, with increasing prevalence especially amongst adolescents and young adults^2,3^. In order to reduce this global burden, early preventative strategies are crucial^4^. With prevention in mind, we focus on the earliest available signs – emotional, behavioural and temperament difficulties arising during the preschool years of early childhood, which have been shown to predict diagnoses of mental illness in later childhood and adolescence^5–7^.

Parental mental health traits, including depression, anxiety and neuroticism, are robust predictors of childhood emotional, behavioural and temperament difficulties^8–10^. Less is known about the possible protective role of parental wellbeing traits on offspring development^11^. Positive affect, life satisfaction and subjective wellbeing go beyond the absence of mental health problems^12^, and can have protective effects on adult mental illness^13^. To gain a more complete picture, we explore the intergenerational transmission of parental wellbeing traits (positive affect, life satisfaction), alongside the transmission of parent mental health traits (anxiety, depression, and neuroticism).

Parental mental health and wellbeing traits can influence offspring outcomes through both genetic and environmental pathways, which can be disentangled using genetically informed family designs. Here we employ a trio-polygenic score (PGS) design^14^ to estimate two possible pathways (see Figure 1). The first pathway “genetic transmission” occurs when the association between parental genotype and child outcome is mediated by child genotype. For example, parents with high genetic predisposition to depression will pass on some of this genetic predisposition, which may manifest in their preschool-aged offspring as emotional, behavioural and/or temperament difficulties. The second pathway “genetic nurture” occurs when the parental genetic predisposition influences child outcomes independently from the child’s genotype, such as through the rearing environments created by the parents. For example, a parent’s genetic predisposition to depression may elevate their own depression risk, which in turn could influence the family environment for example through parenting styles, lifestyles and home organisation. These environmental factors could influence emotional, behavioural and temperamental difficulties in the offspring, over and above genetic transmission^15^. The trio-PGS design allows us to estimate the effects of direct genetic transmission free from confounding by genetic nurture effects, assortative mating and population stratification. While this design also enables interpretable estimates of genetic nurture effects, it is important to acknowledge that these effects may still be confounded by other factors such as rating biases, assortative mating and population structure^16,17^

**Figure 1.**
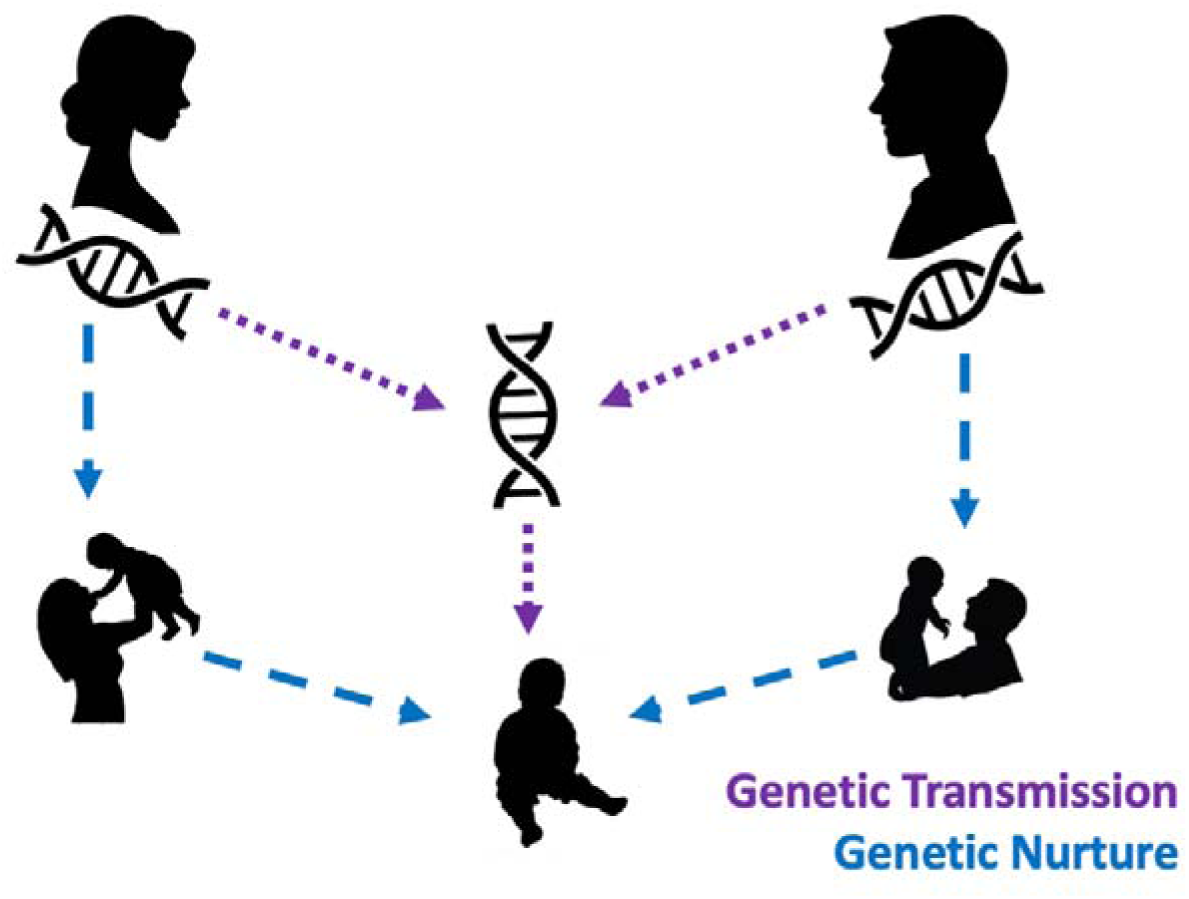
Schematic showing the partitioning of intergenerational transmission into direct genetic transmission effects (through the child’s genotype) in the purple dotted line, and genetic nurture effects (through the parental environments) shown in the blue dashed line.

Genetic nurture can be disentangled from genetic transmission using family-based designs (e.g., adoption, classical twin and children-of-twins designs) or using directly measured genotypes^18^. There is strong evidence across these different study designs that genetic transmission plays a substantial role in offspring mental health^19^ but this is not always consistently found for emotional and behavioural difficulties in the preschool years^20^. The evidence for genetic nurture effects on emotional and behavioural difficulties in preschool children is similarly inconsistent. A meta-analysis across multiple cohorts (including MoBa) found evidence for genetic nurture effects of parental genetic predisposition to neuroticism on offspring emotional difficulties at age 3-years^21^. Whereas, a recent investigation in the Millenium Cohort Study found little evidence for genetic nurture effects from parental genetic predisposition to a range of neuropsychiatric traits on offspring emotional problems at ages 3 and 5-years^20^.

We used a trio-PGS design to explore the intergenerational transmission of mental health and wellbeing traits from parents to preschool-aged offspring. We extend the previous literature to look at the early emergence of emotional, behavioural and temperament difficulties at age 1.5-years, to look at effects on change in these difficulties over time until age 5 years, and to include wellbeing traits as well as mental health traits in the parents.

## Methods

### Sample

We used data from the Norwegian Mother, Father and Child cohort study (MoBa), a population-based pregnancy cohort study^22^. Participants were recruited from across Norway from 1999-2008. Women consented to participate in 41% of pregnancies. The cohort includes approximately 114,500 children, 95,200 mothers and 75,200 fathers. This study is based on version 12 of the quality-assured data files released for research in Jan 2019. Blood samples were obtained from both parents during pregnancy and from mothers and children (umbilical cord) at birth^23^. Quality-control (QC), phasing, imputation, and post-imputation QC of the genotype data is described elsewhere^24^. We had 43,036 mother-father-child trios of European ancestry with complete genetic data and at least one child outcome measure (Table S1 for further sample size information).

### Polygenic scores for parental mental health

We generated PGS for each mental health and wellbeing trait as the weighted sum of risk alleles using PRSice2^25^, restricting to minor allele frequency > 0.01 and independent variants at linkage disequilibrium r^2^=0.1 within 250 kilobases. PGS were regressed on the first 20 principal components (PC) of population structure, genotyping batch and imputation batch. Initially, PGS were generated at 11 p-value thresholds from 5 × 10^-8^ to 1. To avoid multiple testing burden, we used the PGS-PCA method, which takes the first PC from a principal component analysis of PGS at each of the p-value thresholds^26^. PGS generation was done for parents and offspring together, using the same genetic variants.

We used genome-wide association study (GWAS) summary statistics (independent from MoBa) to generate the PGS for depression^27^, anxiety^28^, neuroticism^29^, wellbeing, positive affect, and life satisfaction^30^. GWAS were restricted to individuals of European ancestry for similarity with the genotyped MoBa sample (further details in Note S1).

### Offspring Outcomes

We explored three offspring outcomes: 1) emotional difficulties (anxious and depressive behaviour), 2) behavioural difficulties (inattention, hyperactivity, disruptive behaviour) measured with corresponding subscales of the Child Behaviour Checklist (CBCL)^31^, and 3) irritable temperament measured using the emotionality subscale of the Emotionality, Activity and Shyness Temperament Questionnaire (EAS)^32^. Irritability is a transdiagnostic symptom, seen in both the characterisation of externalising disorders including oppositional defiance disorder and ADHD, as well as internalising disorders including depression and anxiety^33,34^. All scales were mother-reported at ages 1.5-, 3-, and 5-years (Note S2).

### Statistical Analysis

First, we estimated the phenotypic associations between self-reported parental mental health and wellbeing traits on offspring outcomes to ensure associations were present in the MoBa cohort (Note S4). As PGS validation, we checked correlations between PGS were as expected (e.g. large positive association between PGS depression and PGS anxiety) as well as the expected significant associations between each PGS and its respective parental mental health and wellbeing trait (Note S5).

To estimate the pathways of intergenerational transmission (Figure 1) in our trio-PGS analysis, we conducted a mediation model (Figure 2), dividing the total genetic effect into the relative contributions of genetic nurture and genetic transmission. The genetic transmission effect is equivalent to the effect of the parent’s genotype that is mediated by the child’s genotype (i.e. the extent to which inherited genetic variants influence the child’s outcomes) (dotted lines, Figures 1-2). The genetic nurture effect is estimated as the residual association between parental PGS and child’s emotional and behavioural difficulties, assumed to associate via the genetically predicted environments that the parent provides (dashed lines, Figures 1-2). This mediation model is statistically equivalent to trio-PGS models regressing offspring outcomes jointly onto their own and both their parents’ PGS (e.g., ^35^).

**Figure 2.**
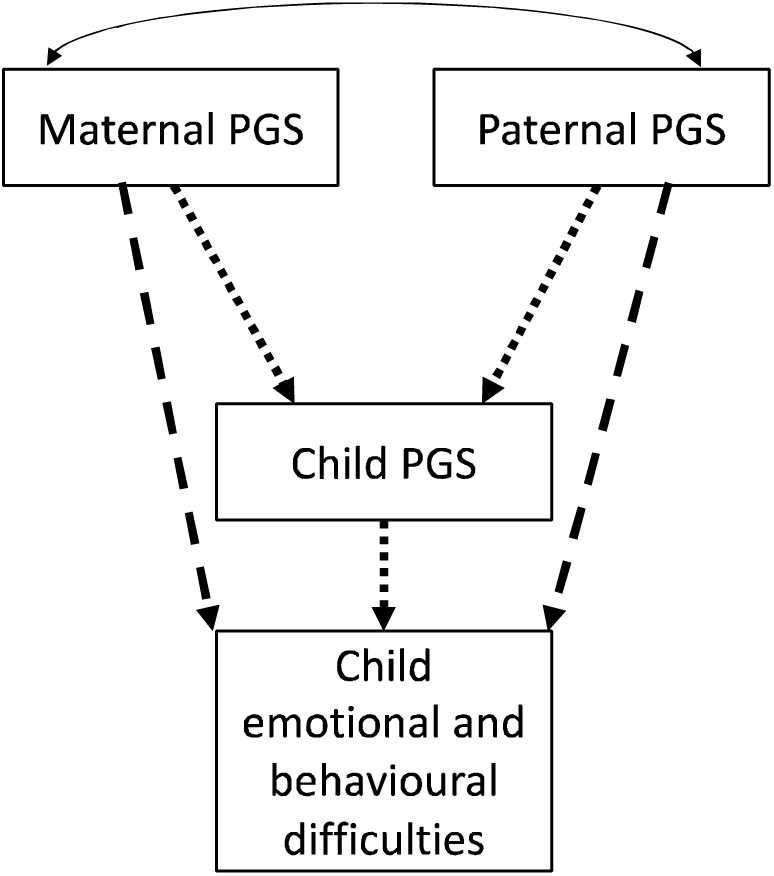
Trio-polygenic score model for parental mental health on offspring emotional, behavioural and temperament difficulties (dotted arrows represent genetic transmission effects, dashed arrows represent genetic nurture effects, double headed arrow represents covariance between maternal and paternal PGS). PGS = polygenic score.

To investigate associations with both stability and change in offspring outcomes, we parameterised each in terms of a latent growth process (details in Note 3). This meant that, for each outcome, we included both a latent intercept – corresponding to variance present at initial assessment (1.5-years) and persisting across development – and a latent slope – corresponding to variability in linear change in outcomes between 1.5- and 5-years – as outcomes in the trio-PGS models. As secondary outcomes, we estimated genetic transmission and genetic nurture effects on observed traits at each individual time point.

We estimated mediation models using structural equation modelling in the *lavaan* package^36^ (v0.6.7) in R^37^ (v4.0.3). Child sex was included as a time-invariant covariate. Our primary models included individuals in the full sample with data available on at least one relevant measure (genotype data for parents; n=53,032 fathers, n=77,216 mothers, and either genotype, outcome or covariate data for offspring; n=113,276 children) using full information maximum likelihood (FIML) estimation to account for missing data. We clustered on maternal identifier to account for mothers reporting on multiple offspring (siblings). Although this clustering partially accounts for relatedness in the sample, it does not account for all possible relatedness. Therefore, we compared the results from the full sample with those from a sample restricted to unrelated complete genotyped trios only (Note S6 for this and other sensitivity tests including assortative mating).

## Results

We showed that PGS were valid predictors of observed parental traits (Tables S3-S8). Furthermore, observed mental health and wellbeing traits in both parents were strongly associated with emotional, behavioural and temperament difficulties in their offspring (Table S2), justifying exploration of intergenerational transmission in this cohort.

Overall, genetic transmission effects were small, often with confidence intervals crossing the null (Figure 3, Table S9). Estimates of genetic nurture were less precise (Figure 3) but tended to explain a greater percentage of variance for most offspring outcomes (Figure 4). Across most offspring outcomes, we saw evidence for genetic nurture effects (Figure 3). Comparisons of model fit revealed significantly larger genetic nurture effects from wellbeing traits than mental health traits for change in emotional difficulties, stability and change in behavioural difficulties (Table S15). As an example, genetic nurture effects of maternal depression PGS on initial levels of behavioural difficulties = 0.013 (95% CI: −0.011, 0.037), p=0.30, compared with genetic nurture effects of maternal wellbeing PGS on the same outcome = −0.050 (95% CI: −0.074, −0.026), p<0.001. There was a trend towards larger maternal genetic nurture effects than paternal. We formally tested whether this difference was significant my comparing model fit. Significant differences are depicted in Figure 3 and include significantly larger maternal than paternal genetic nurture effects for anxiety PGS on behavioural stability, irritability stability and change, and significantly larger maternal than paternal genetic nurture effects for positive affect and life satisfaction PGS on stability of emotional difficulties and change in behavioural difficulties (Figure 3, Table S14).

**Figure 3.**
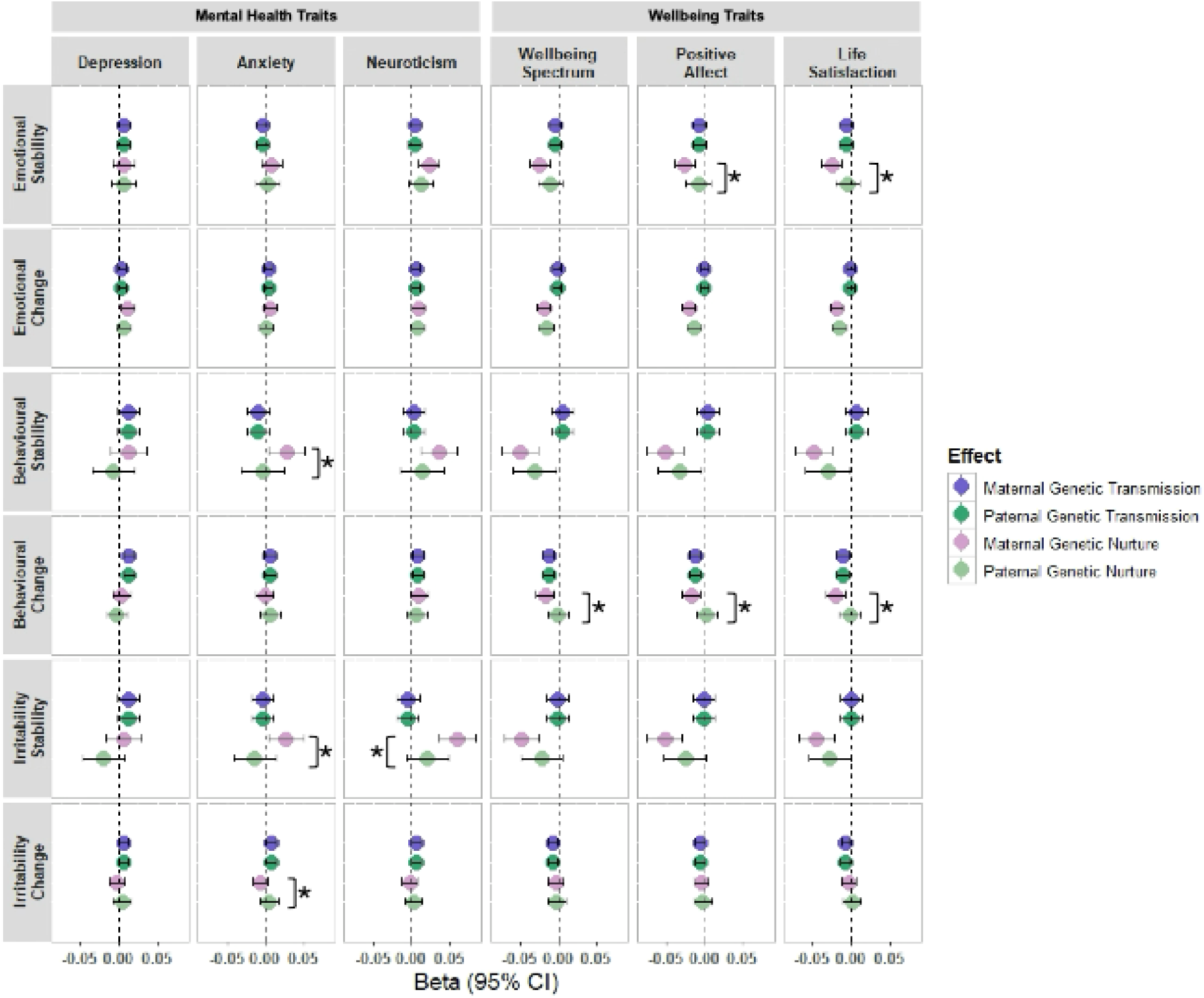
The association between parental polygenic risk for mental health traits on offspring emotional and behavioural difficulties. Genetic effects are decomposed into that which can be accounted for by genetic transmission directly from parent to offspring, and that which can be accounted for by the genetically influenced environments provided by the parents (genetic nurture). * = influence of maternal and paternal genetic nurture is significantly different, as determined by a significant (p<0.05) reduction in model fit after constraining maternal and paternal genetic nurture effects to be the same.

**Figure 4.**
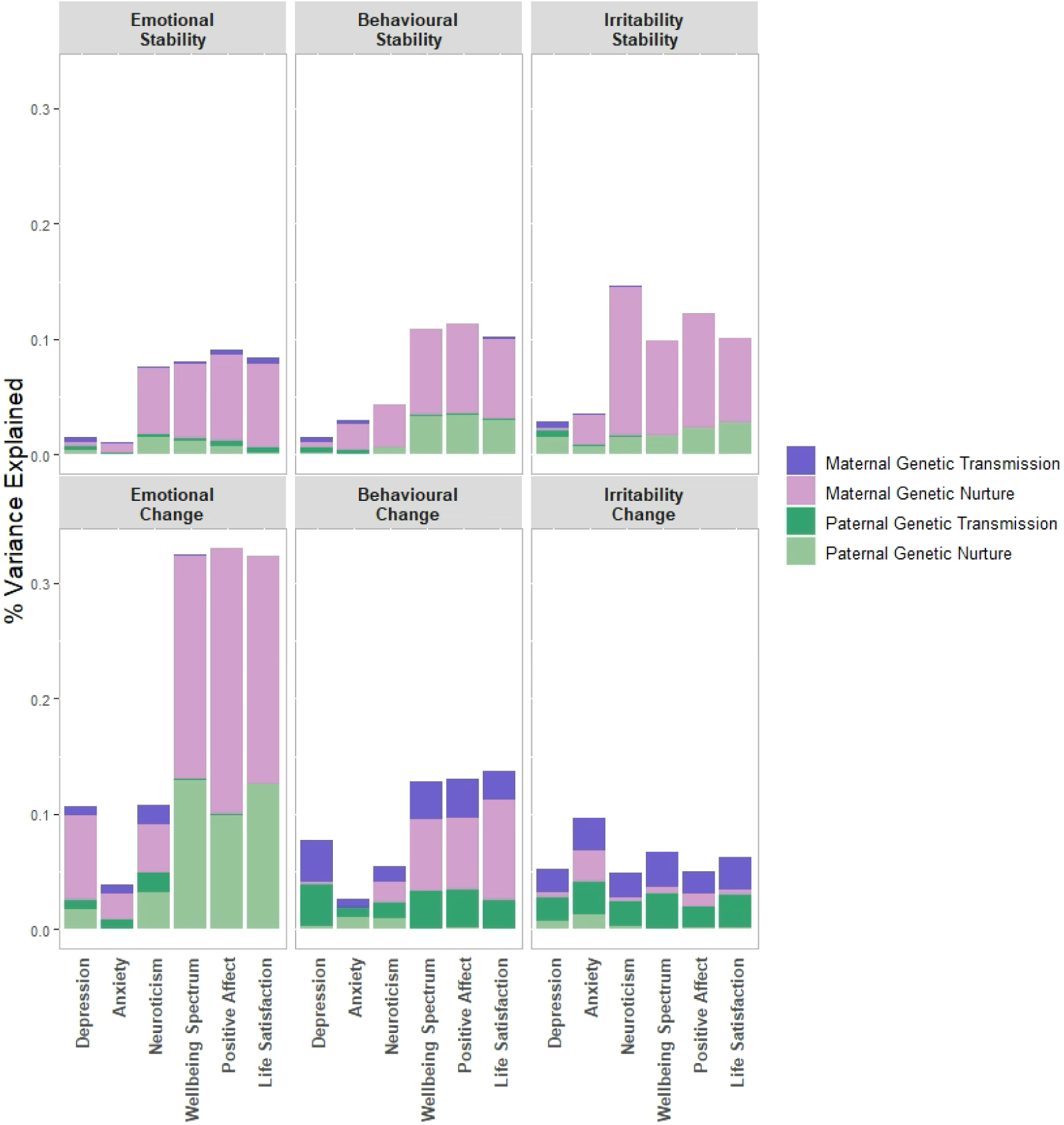
The relative contribution of genetic transmission and genetic nurture to percentage of explained variance in stability and change of emotional and behavioural difficulties.

Overall, stronger genetic transmission and genetic nurture effects were observed for offspring trait stability (intercept) compared to the effect on change in offspring traits (slope) between ages 1.5- to 5-years (Figure 3). Exceptions were wellbeing traits PGS predicting developmental reductions in emotional difficulties, driven by both maternal and paternal genetic nurture effects (e.g. maternal wellbeing PGS = −0.019, 95%CI= −0.027, −0.010, p<0.001) and maternal wellbeing trait PGS associated with a reduction of behavioural difficulties over time (e.g. maternal wellbeing PGS = −0.017, 95%CI= −0.029, −0.005, p=0.005).

Results for secondary, univariate models were relatively consistent (Figures S1-S3; Table S10). Sensitivity analyses found that results were consistent when restricting to unrelated complete genotyped trios (Figures S4-S5; Table S11), using unweighted scores (Figures S6-S7; Table S12), and there was little evidence of assortative mating (Table S13).

## Discussion

We leveraged large-scale trio data from the MoBa cohort to examine the intergenerational transmission of parental mental health and wellbeing traits on offspring’s emotional, behavioural and temperament difficulties in the preschool years. Our results suggest that genetic transmission effects are generally small at this young age, with confidence intervals often encompassing null values. Our findings support the relative importance of genetic nurture effects in early childhood. However, our estimates of genetic nurture effects can still be confounded by other factors including rating biases and population structure^16,17^. Differentiating pathways of intergenerational transmission has implications for intervention. If intergenerational associations are explained purely by genetic transmission, then interventions focusing on parent mental health are less likely to influence child mental health. However, if intergenerational transmission is environmentally mediated, then interventions improving parent mental health or other environmental mechanisms are likely to also benefit offspring mental health.

Our study contributes novel insights by examining the intergenerational transmission of wellbeing traits: wellbeing spectrum, life satisfaction, and positive affect. We observed larger genetic nurture effects from wellbeing trait PGS than mental health trait PGS for emotional and behavioural difficulties. Our findings are consistent with previous literature which suggests that positive childhood environments, influenced by parental wellbeing, such as supportive and nurturing parenting, may act as protective factors against the development of emotional and behavioural difficulties in children. Examples of possible positive parenting environments include: parental knowledge of offspring whereabouts, parental warmth, and strong parent– child relationship, which have previously been associated with lower emotional and behavioural difficulties^38–41^. Future research should investigate these (and other) positive environments possibly on the genetic nurture pathway to offspring mental health, in order to foster resilience, even in the presence of genetic transmission of mental illness.

Overall, the evidence for genetic transmission effects was small. We know that parents and offspring share 50% of their genetic material, and that both parental and childhood mental health traits are heritable^42^, highly polygenic and genetically correlated^43^. One possible explanation for low estimates of genetic transmission could be that our model uses PGS for *adulthood* mental health traits to predict offspring emotional and behavioural difficulties in early childhood. This makes theoretical sense given our interest in transmission from adult parents to child offspring, and phenotypic profiles of mental health difficulties are distinct, presenting differently across age groups. However, PGS often lack predictive power across different contexts. Power calculations in a similar size sample of the MoBa cohort revealed 80% power to detect genetic transmission effects of 0.02^44^. For the most part, genetic transmission effects observed here were smaller, suggesting we might have been underpowered to detect small effects. Alternatively, genetic transmission effects might simply be smaller in preschool years. There is evidence to suggest that estimates of heritability increase as we age for various traits^45,46^ including for emotional and behavioural difficulties^47^, and the contribution of specific genetic variants changes over age. Supporting this, a recent exploration in MCS found evidence that genetic transmission effects on emotional difficulties increased from childhood to adolescence, with a similar lack of evidence for genetic transmission effects at ages 3- and 5-years^20^.

After accounting for these small effects of genetic transmission, what remains of the total genetic effect could come from several sources. Of primary interest are the genetic nurture effects, mediated through parental environments. However, it is important to note that our model estimates of this pathway can be confounded by other factors such as rating biases, population structure, and assortative mating^16,17^. We have taken steps to mitigate these biases where possible, adjusting for 20 principal components of population structure and conducting sensitivity analyses for assortative mating. However, we are limited in only having maternal reports of offspring outcomes. Maternal genetic predisposition to mental health traits could influence reporting. This pathway is independent of child genotype and would therefore falsely inflate genetic nurture effects. This could explain the presence of larger maternal compared to paternal genetic nurture effects for several outcomes. Stronger evidence for maternal genetic nurture effects is consistent with previous studies in the MoBa cohort, finding that maternal effects of neuroticism on offspring depression and anxiety persisted after accounting for genetic transmission and assortative mating, whereas paternal effects were greatly attenutated^48^.

However, without reports from father or independent raters, we cannot conclude whether estimates are inflated by maternal reports, or for other reasons such as due to the mother on average spending more time with offspring. For example, in Norway during the recruitment period of MoBa, fathers did on average a 35-38% share of the childcare^49^. However, we do still detect evidence for paternal genetic nurture effects, suggesting the effects observed are not due to maternal reporting bias alone.

Furthermore, evidence for a predominance of genetic nurture effects in early childhood is consistent with previous literature from multiple study designs. For example, there is evidence from children of twin and adoption designs that the effects of parental depressive symptoms on offspring emotional and behavioural problems persist after adjusting for genetic transmission^50^.

Furthermore, the classical twin design often finds a contribution of shared environment (C) in early childhood, which reduces over age^51^. Developmentally, this suggests that the contribution of parental environments is likely to be greatest in the early years of life^19^. Our findings are consistent with genetic investigations of mental health and neurodevelopment in young childhood, showing evidence for genetic nurture effects of parental neuroticism on offspring emotional difficulties at age 3-years^21^ and of whole measured genotypes on neurodevelopmental traits at age 3-years^52^. However, one study found no evidence for genetic nurture effects at 3- or 5-years using transmitted and non-transmitted parental psychiatric genetic predisposition on youth emotional problems in a smaller UK population (n<3,000)^20^.

However, their smaller sample had less power to detect genetic nurture effects. In mid-childhood, the pattern of results continues to appear inconsistent. Some studies find evidence for genetic nurture effects of parental neuropsychiatric traits on emotional and behavioural offspring outcomes at 8-years^15^, and for genetic nurture effects from whole genotypes on childhood depressive symptoms at 8-years^53^ and neurodevelopmental traits at 8-years^54^. Other studies find no strong evidence for significant genetic nurture effects on anxiety symptoms at 8-years^53^ nor on depressive, disruptive, or ADHD traits at 8-years^55^ nor conduct problems at 8-years^44^. Several of these studies had smaller samples and were perhaps less well powered to detect genetic nurture effects than more recent investigations^52^. Alternatively, genetic nurture effects on emotional, behavioural and temperament difficulties might be more developmentally limited to younger childhood and less pronounced by age 8-years, suggesting that prevention and early intervention strategies may have to differ depending on age.

### Strengths and limitations

There were several strengths to this study. First, we leveraged a large sample of mother-father-child trios from the MoBa cohort to disentangle pathways of intergenerational transmission. Second, our statistical models accounted for missingness and relatedness, showing relative consistency with complete genotyped trios. Third, we included measures of wellbeing traits, which allow for investigation of resilience and protective effects. Fourth, we employed broad indicators of emotional and behavioural difficulties, including irritable temperaments, recognizing the nonspecific, developmentally dependent nature of these difficulties in childhood^56,57^. Finally, rather than a single time-point measure of child outcomes, we investigated trajectories spanning 1.5- to 5-years, providing a more comprehensive understanding of how intergenerational transmission influences change over age.

However, there are several limitations. First, we cannot rule out possible bias due to assortative mating or population stratification^18^ despite steps taken to mitigate this. Second, we only had mother-reports of offspring outcomes which could inflate maternal estimates. Future studies should replicate using father reports or objective assessments. Third, the genotyped sample of MoBa currently only contains individuals of European ancestry and GWAS of parental mental health traits were derived in European Ancestry Populations, therefore results might not generalise to individuals of other ancestries. Furthermore, parenting roles in Norway may not generalise to other countries. Finally, strength of conclusions will be improved as more powerful GWAS of mental health and wellbeing traits become available, in particular, those with greater temporal proximity to the age of our child outcomes. Such developmentally specific GWAS may also be better able to capture effects on change.

## Conclusions

In conclusion, this study leveraged family trio data from the MoBa cohort to explore intergenerational transmission from parental mental health and wellbeing traits to preschool emotional, behavioural and temperament difficulties. Understanding the pathways to early-life outcomes is important for preventing future mental illness, as early intervention can significantly improve prognosis^4^. We found evidence for a greater relative contribution of genetic nurture on emotional difficulties, behavioural difficulties and irritability in preschool-aged offspring, and a significantly greater contribution of genetic nurture from wellbeing trait PGS than mental health trait PGS, highlighting the need for positive parenting interventions.

Future work should aim to identify the extent to which estimates of genetic nurture represent environmentally-mediated, causal effects – and which environments in particular are involved – in order to identify suitable targets for intervention testing.

## Ethical Statement

The establishment of MoBa and initial data collection was based on a license from the Norwegian Data Protection Agency and approval from The Regional Committees for Medical and Health Research Ethics. The MoBa cohort is currently regulated by the Norwegian Health Registry Act. The current study was approved by The Regional Committees for Medical and Health Research Ethics (2016/1702).

## Data Availability

All data produced in the present work are contained in the manuscript. The data on which these analyses are based (the MoBa cohort) is available through the cohort upon application.

https://www.fhi.no/en/ch/studies/moba/for-forskere-artikler/research-and-data-access/

## Acknowledgements

The Norwegian Mother, Father and Child Cohort Study is supported by the Norwegian Ministry of Health and Care Services and the Ministry of Education and Research. We are grateful to all the participating families in Norway who take part in this on-going cohort study. This work was performed on the TSD (Tjeneste for Sensitive Data) facilities, owned by the University of Oslo, operated and developed by the TSD service group at the University of Oslo, IT-Department (USIT). The analyses were performed on resources provided by Sigma2 - the National Infrastructure for High Performance Computing and Data Storage in Norway. We thank the Norwegian Institute of Public Health (NIPH) for generating high-quality genomic data. This research is part of the HARVEST collaboration, supported by the Research Council of Norway (#229624). We also thank the NORMENT Centre for providing genotype data, funded by the Research Council of Norway (#223273), South East Norway Health Authorities and Stiftelsen Kristian Gerhard Jebsen. We further thank the Center for Diabetes Research, the University of Bergen for providing genotype data and performing quality control and imputation of the data funded by the ERC AdG project SELECTionPREDISPOSED, Stiftelsen Kristian Gerhard Jebsen, Trond Mohn Foundation, the Research Council of Norway, the Novo Nordisk Foundation, the University of Bergen, and the Western Norway Health Authorities.

